# Early assessment of molecular progression and response by whole-genome circulating tumor DNA in advanced solid tumors

**DOI:** 10.1101/19002550

**Authors:** Andrew A. Davis, Wade T. Iams, David Chan, Michael S. Oh, Robert W. Lentz, Neil Peterman, Alex Robertson, Abhik Shah, Rohith Srivas, Timothy Wilson, Nicole Lambert, Peter George, Becky Wong, Haleigh Wood, Jason Close, Ayse Tezcan, Ken Nesmith, Haluk Tezcan, Young Kwang Chae

## Abstract

Treatment response assessment for patients with advanced solid tumors is complex and existing methods require greater precision. Current guidelines rely on imaging, which has known limitations, including the time required to show a deterministic change in target lesions. Serial changes in whole-genome (WG) circulating tumor DNA (ctDNA) were used to assess response or resistance to treatment early in the treatment course. 96 patients with advanced cancer were prospectively enrolled (91 analyzed and 5 excluded), and blood was collected before and after initiation of a new, systemic treatment. Plasma cell-free DNA libraries were prepared for either WG or WG bisulfite sequencing. Longitudinal changes in the fraction of ctDNA were quantified to retrospectively identify molecular progression (MP) or major molecular response (MMR). Study endpoints were concordance with first follow-up imaging (FFUI) and stratification of progression-free survival (PFS) and overall survival (OS). Patients with MP (n=13) had shorter PFS (median 62d vs. 310d) and OS (255d vs. not reached). Sensitivity for MP to identify clinical progression was 54% and specificity was 100%. MP calls were from samples taken a median of 28d into treatment and 39d before FFUI. Patients with MMR (n=27) had longer PFS and OS compared to those with neither call (n=51). Molecular response assessment can potentially enable early switching to potentially effective therapies, therefore minimizing side effects and costs associated with additional cycles of ineffective treatment. MMR may present a novel endpoint to target to improve long-term patient outcomes.

## Introduction

The current standard of care for clinical evaluation of treatment response for advanced solid tumors is based on physical exams, patient reported symptoms, and periodic radiographic tumor assessments that are compared with baseline scans to determine whether the patient is responding or progressing while on active treatment. However, there are limitations to these approaches as subtle changes in disease are often asymptomatic, and considerable costs, uncertainty, and anxiety can be associated with frequent imaging. In clinical trials, imaging response criteria are standardized to guide evaluation by comparing a baseline scan before treatment initiation with periodic follow-up imaging with pre-specified criteria for response (Eisenhauer et al. 2009; Seymour et al. 2017). These criteria can be limited by the reliability of measurements over time, difficult to measure sites of disease (e.g., bone or pleural effusions), and challenges distinguishing pseudoprogression from true progression (Erasmus et al. 2003; Nishino et al. 2017; Chiou and Burotto 2015). In addition, inter-operator variability in comparing serial scans remains a limitation in response assessment (Kuhl et al. 2019). Therefore, novel methods for monitoring response to treatment are needed given the emergence of new treatment modalities with ongoing questions regarding how best to manage treatment, minimize toxicity, and control costs.

Liquid biopsy assays in cancer patients may analyze circulating cell-free DNA (cfDNA), circulating tumor cells (CTCs), RNA, exosomes, or proteins (Wan et al. 2017; Siravegna et al. 2017). ctDNA likely originates from cancer cells undergoing apoptosis, necrosis, or potential active mechanisms involving nucleic acid secretion to facilitate metastasis and gene expression at distant sites (Heitzer et al. 2015). The amount of ctDNA correlates with more advanced stages of disease and is also affected by tumor type, origin, location of metastasis, and treatment (Chen et al. 2016; Merker et al. 2018). There are several distinguishing features between ctDNA and non-tumor cfDNA. Specifically, as compared with cfDNA, ctDNA contains tumor-specific somatic point mutations, structural variations, shorter fragment lengths, biased fragment start and end positions, and changes in epigenetic patterns (Murtaza et al. 2013; Chan, Jiang, Zheng, et al. 2013; Mouliere et al. 2011; Jiang et al. 2015; Underhill et al. 2016; Snyder et al. 2016; Jiang et al. 2018; Sun et al. 2015).

Copy number aberrations (CNA), which consist of either deletions or duplications of portions of the genome, are a common form of structural variation that are observed in patients with advanced disease at various sites across the genome (Beroukhim et al. 2010). Prior studies have demonstrated that CNAs can be detected in cfDNA from patients by low-pass next-generation sequencing (NGS) with CNAs detected at a higher rate in patients with advanced disease (Mouliere et al. 2018; Adalsteinsson et al. 2017). To date, changes in CNAs over time in advanced cancer patients remain understudied.

Global hypomethylation is a hallmark of tumor genomes (Sun et al. 2015; Hon et al. 2012), and an increase in global methylation levels in cfDNA might be associated with non-progression as it would indicate a decreased proportion of ctDNA. Importantly, the epigenetic patterns observed in tumors, including overall global hypomethylation can be detected in ctDNA and could therefore have potential for tracking patients over time (Chan, Jiang, Chan, et al. 2013; Usadel et al. 2002).

Recently, there has been significant interest in evaluating the potential clinical utility of ctDNA in advanced solid tumors. In the adjuvant setting, residual ctDNA after surgery (e.g., minimal residual disease) has been associated with disease recurrence across multiple tumor types (Pantel and Alix-Panabières 2019; Abbosh et al. 2018; Chae and Oh 2019; Merker et al. 2018; Rossi and Ignatiadis 2019). In advanced disease, the most well validated clinical use is to identify driver mutations with known drug targets (Lanman et al. 2015; Diaz and Bardelli 2014; Reckamp et al. 2016; Juric et al. 2019). Additionally, resistance mutations have been identified in the blood with the goal of guiding clinical management (Fribbens et al. 2016). While prior studies have evaluated the potential role of tracking ctDNA for tumor response assessment in several solid tumor types, typically tracking mutant allele frequencies or acquired resistance mutations, the clinical utility for routine assessment has not yet been established (Diehl et al. 2008; Dawson et al. 2013; Murtaza et al. 2013; Tie et al. 2015; Pécuchet et al. 2016; Hrebien et al. 2019; Merker et al. 2018).

Here, in a prospectively enrolled, advanced stage, pan-cancer cohort, we performed WG cfDNA analysis to use ctDNA as a molecular marker for response assessment earlier in the treatment course compared with routine clinical and radiographic assessment of disease. In contrast to analyzing particular genes, our approach utilized CNAs and fragmentation patterns across the genome, a technique with broad potential clinical applications across multiple tumor types. Furthermore, for a subset of patients, we performed bisulfite conversion as part of the assay, which provided insight into genome-wide methylation changes. We hypothesized that early changes in cancer-associated signals in the blood would be predictive of response status at the time of FFUI and that the magnitude of the dynamic change in signal would provide longer-term prognostic information across a variety of solid tumor types and treatments.

## Patients and Methods

### Study subjects

The study sample here represented a subset of a currently accruing longitudinal observational study. From May 2017 to December 2018, participants (age >18 years) were prospectively enrolled from five oncology centers in the USA (TMPN - Cancer Care, Redondo Beach, CA; Scripps - California Cancer Associates, San Diego, CA; Sharp Memorial Hospital, San Diego, CA; Summit Cancer Centers, Post Falls, ID; Robert H. Lurie Comprehensive Cancer Center of Northwestern University, Chicago, IL) and followed through September 2019 (Table 1). Though five centers were enrolled in the study, only participants from four sites were included in this analysis because one site only enrolled sarcoma patients, which were held for future studies. Eighty percent of the participants in this analysis came from one center (Supp. Table S1). Eligibility criteria were:

1. Diagnosis of a non-hematologic and surgically unresectable advanced tumor (stage III or higher) at presentation
2. Commencement of a new systemic treatment regimen of the physician’s choice
3. Presence of either measurable or evaluable disease by imaging

**Table 1.** Participant characteristics. May 2017 - September 2019.

|  | **Median (Min-Max)** | **N= 91 (%)** |
| --- | --- | --- |
| **Age** | 70 (30-89) |  |
| **Sex** |  |  |
| Female |  | 50 (55) |
| Male |  | 41 (45) |
| **Cancer type** |  |  |
| Lung |  | 40 (44) |
| Breast |  | 24 (26) |
| GI |  | 14 (15) |
| GU |  | 6 (7) |
| Melanoma |  | 6 (7) |
| Sarcoma |  | 1 (1) |
| **Treatment types** |  |  |
| Chemotherapy |  | 32 (35) |
| Chemotherapy, Antibody |  | 10 (11) |
| Immunotherapy |  | 24 (26) |
| Immunotherapy, Chemotherapy |  | 9 (10) |
| Immunotherapy, HDACi |  | 1 (1) |
| Endocrine |  | 4 (4) |
| Endocrine, CDK4/6i |  | 6 (7) |
| Targeted alone |  | 5 (5) |
| **Lines of therapy** |  |  |
| 1 |  | 48 (53) |
| 2 |  | 23 (25) |
| 3+ |  | 20 (22) |
| **Timing (days since treatment start)** |  |  |
| T1 | 21 (14-40) | 85 (93) |
| T2 | 42 (37-84) | 66* (72) |
| First follow-up | 69 (26-208) |  |
| Last follow-up | 384 (60-754) |  |
| **Protocol** |  |  |
| WGS |  | 53 (58) |
| WGBS |  | 38** (42) |

\*60 had both post-treatment time points. \*\*In addition, 13 of the participants analyzed with WGS were also analyzed with WGBS.

To be included in this cohort, the participants needed to have venous blood samples from at least two time points: a baseline (T0, before treatment initiation) and another one before cycle 2 (T1) and/or cycle 3 (T2, Fig. 1A). This timing for our assay, in line with the standard clinical visits was designed to assess whether it could be of use if integrated into current practice without adding additional appointments. The timing in our study was in line with prior work optimizing the collection timing of liquid biopsy samples for use as a surrogate for PFS (Hrebien et al. 2019). The study reported here was conducted in accordance with the Declaration of Helsinki and approved by Northwestern University, Sharp Memorial Hospital and Western Institutional Review Boards. An informed consent was obtained from each patient prior to participation in the study.

**Figure 1.**
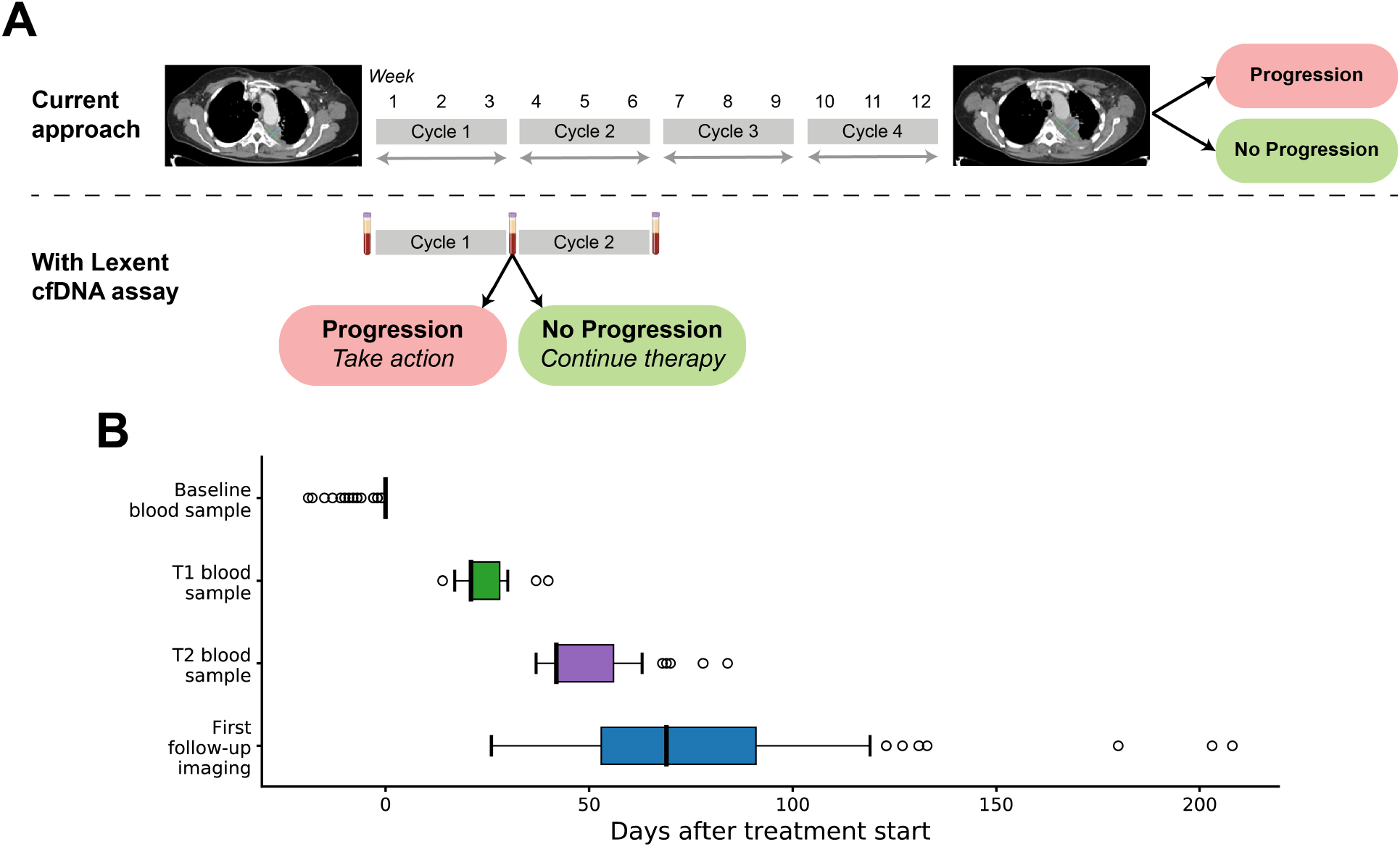
Overview of the clinical setting. **A**. Diagram comparing radiographic response assessment and the potential use of cfDNA to assess molecular response. **B**. Timing of imaging and blood collections for patients in the study.

### Evaluation of response status

Participants were radiologically assessed at baseline and again at first follow-up as determined per standard of care routine clinical assessment. The primary endpoint of the study was evidence of radiographic progression as determined by Response Evaluation Criteria in Solid Tumors (RECIST) version 1.1 (Eisenhauer et al. 2009) or clinical response evaluation. Measurable disease changes by imaging were interpreted by an independent radiologist, who was blinded to the assessment of molecular response. Radiographic assessments were categorized as progressive disease (PD) or non-progressive disease (nonPD) which included both partial response (PR) and stable disease (SD). When RECIST outcome could not be ascertained due to either non-evaluable or missing imaging scans, clinical response evaluation was used with PD defined based on the physician’s outcome assessment before a treatment change.

OS was defined as the time from the start of treatment to death due to any cause. PFS was defined as the time from the start of treatment to first documentation of PD, or death due to any cause, whichever occurred first. Patients last known to be alive and progression-free were censored at the date of last follow-up. Patients were considered as lost to follow-up if they were no longer part of the study and their status after FFUI was unknown.

### Sample preparation

At each time point, 10 mL of whole blood was collected in Streck Cell-Free DNA BCT. Plasma was separated via centrifugation at 1600 x g for 15 minutes followed by 2500 x g for 10 minutes within 7 days from the time of collection. cfDNA was extracted from plasma using the Qiagen QIAmp MinElute ccfDNA kit and stored at −20C until library preparation. For each patient, libraries were prepared using the KAPA HyperPrep library prep kit for whole genome sequencing (WGS; n=53 patients) or the Nugen Ovation Ultralow Methyl-Seq whole genome bisulfite sequencing kit (WGBS; n=38 patients). Average input cfDNA into the library preparation was 20 ng. Libraries were sequenced on the Illumina HiSeq X to an average depth of 20X (range 6-29X).

### Bioinformatics methods

Reads were aligned to the human genome (GRCh37) with a custom bioinformatics pipeline based on BWA (Li and Durbin 2009), sambamba (Tarasov et al. 2015), and samtools (Li et al. 2009). WGBS libraries were processed with an adapted pipeline to align sequencing data to a bisulfite-converted human genome (Krueger and Andrews 2011). Reads were then de-duplicated and GC biases were corrected using the deepTools software package (Ramírez et al. 2014). Sequencing quality was validated using Picard (Broad Institute 2019) to assess mapping efficiency, GC bias, and duplication rate, as well as MethylDackel (Ryan 2017) for WGBS libraries to measure bisulfite conversion efficiency.

Tumor fraction ratio (TFR) was measured to assess changes in ctDNA using CNAs and local changes in cfDNA fragment length, both assessed from sequencing data. CNAs were detected using a pipeline based on ichorCNA (Adalsteinsson et al. 2017) and custom algorithms. Normalized fragment length was computed by normalizing by library and genomic location. Background signals for CNA and fragment length were established for each sequencing protocol using healthy normal samples taken from 44 individuals with no current or prior diagnosis of any malignancy (Supp. Table S2). In order to maximize sensitivity of CNAs while preventing false positive detections, Spearman’s rank correlation between local mean fragment length and copy number was used as a disqualifier of CNA call sets (Hellwig et al. 2018; Mouliere et al. 2018).

To assess changes in ctDNA over time, direct comparison of the CNA-derived estimate of absolute tumor fraction between a patient’s time points was not always reliable because there was ambiguity in which read depth levels correspond to which structural events. For example, two regions might be called as either a neutral region and a duplicated region, or a heterozygous deletion and a neutral region, in a highly mutated tumor where there was an ambiguous neutral level. To circumvent this, CNAs detected at multiple time points were compared longitudinally with a linear model to quantify TFR. In order to determine confident calls, measured changes were compared to a simulated background model and required to exceed a Z-score threshold of 3. No longitudinal comparisons of samples from 44 healthy participants (Supp. Table S1) showed a significant change in TFR (e.g. Supp. Fig. S1). Cases that showed a confident increase in tumor fraction indicated by TFR greater than 1 at either time point were retrospectively classified as molecular progression (MP). A confident increase was defined to be a TFR between a follow up time point and the baseline of 1 plus a confidence interval equivalent to three standard deviations based on the variation in depth in the copy number altered regions in an unaffected participant library. Major molecular response (MMR) was defined as a TFR < 0.1, an arbitrary cutoff (1 log reduction) selected prior to PFS and OS assessment.

### Statistical Analysis

For the purposes of calculating sensitivity and specificity, true positives were defined as cases where the assay showed a molecular progression which were also evaluated as PD either clinically or radiographically at FFUI; true negatives were cases with no MP and clinical or radiographic nonPD at FFUI. False positives and false negatives were cases where molecular response assessment disagreed with the clinical or radiographic assessment at FFUI of PD or nonPD, respectively. Confidence intervals on these metrics were computed with the Wilson’s score interval method.

Survival curves were generated using the Kaplan-Meier method for PFS and OS. The Cox proportional hazards model was used to analyze the association of assay results and clinical covariates to PFS and OS. The proportional hazards assumption was evaluated with a score test on Schoenfeld residuals (Grambsch and Therneau 1994). Differences in survival were assessed with the Wald test. For two groups in the subset analysis, the Cox regression did not converge due to the small number of events in these subgroups (Immunotherapy PFS and Endocrine/Targeted therapy OS), so a ridge penalty was applied to assay results (Verweij and Van Houwelingen 1994). For multivariable analysis, line of therapy was grouped into treatment naive (1st line of therapy) and previously treated (2nd or greater line). All P values were two-sided. Statistical analyses of survival were performed with the R survival package version 2.41-3 and other statistical analyses were determined using the python scipy package version 1.1.0.

## Results

### Patient characteristics

A total of 96 patients with advanced cancer, who met the inclusion criteria of the study and had at least FFUI, were sequenced for the analysis (Supp. Fig. S2). Baseline blood samples failed sequencing for five participants and were therefore excluded. The remaining 91 patients with NGS and clinical outcome data were included in this analysis. The median age was 70 years and 55% were females (Table 1). About half of the participants received a first-line therapy (53%) and 25% received a second-line therapy. The majority of patients had lung cancer (44%) or breast cancer (26%), with the remainder of the cohort representing gastrointestinal cancers (GI, 15%, specific types in Supp. Table S3), genitourinary cancers (GU, 7%), melanoma (7%) and sarcoma (1%). During the study, 46% of all participants received chemotherapy (+/- antibody treatment, i.e. trastuzumab, pertuzumab, panitumumab, ramucirumab, bevacizumab, olaratumab) and 37% received immunotherapy with or without chemotherapy or a histone deacetylase inhibitor (Table 1, Supp. Table S4). FFUI occurred a median of 69 days after treatment start (Fig. 1B, min=26 days, max=208 days). Three participants received an outcome assignment based on clinical evaluation. Of these three participants, one had a non-evaluable imaging study and was assessed as clinical nonPD at week-12, and two did not have an imaging study prior to treatment change, and these patients were assessed as clinical PD at week-9 and week-19 by the treating physician. The median follow-up time of the full cohort was 384 days (min=60 days, max=754 days).

Baseline blood samples were collected prior to treatment start for all patients (Fig. 1B, median=0 days after treatment start, earliest 19 days before treatment start). One or two post-treatment samples were collected for each patient, with T1 collected prior to the second cycle of therapy at a median of 21 days after treatment start (n=85, min=14 days, max=40 days) and T2 collected prior to the third cycle of therapy at a median of 42 days (n=66, min=37 days, max=84 days). Most blood samples were from before FFUI; however, there were some exceptions with three T1 samples taken the same day or later (median 44 days before FFUI) and 12 T2 samples taken the same day as or later (median 26 days before FFUI). Both post-treatment samples were collected for 60 patients.

### Serial measurements of ctDNA show rapid changes early on treatment

We assessed changes in tumor fraction using WG analysis to quantify the TFR between baseline and post-treatment samples. Substantial changes in TFR were observed early after treatment initiation (Fig. 2). Patient LS030178 is included as an example of a rapid increase in TFR to 2.4 at T1 following the first cycle of therapy, indicating a major increase from baseline, followed by an even greater increase at T2 (Fig. 2A-D). This patient had a somatic gain of the long arm of chromosome 1 (1q), which has been shown to be one of the most common arm-level aberrations in breast cancer (Berger et al. 2018). Additionally, the strong pattern of CNAs was corroborated by the fragmentation pattern (Fig. 2B-C). Conversely, patient LS030093 showed a decrease in TFR at T1 and then a larger decrease at T2, indicating response to treatment (Fig. 2E).

**Figure 2.**
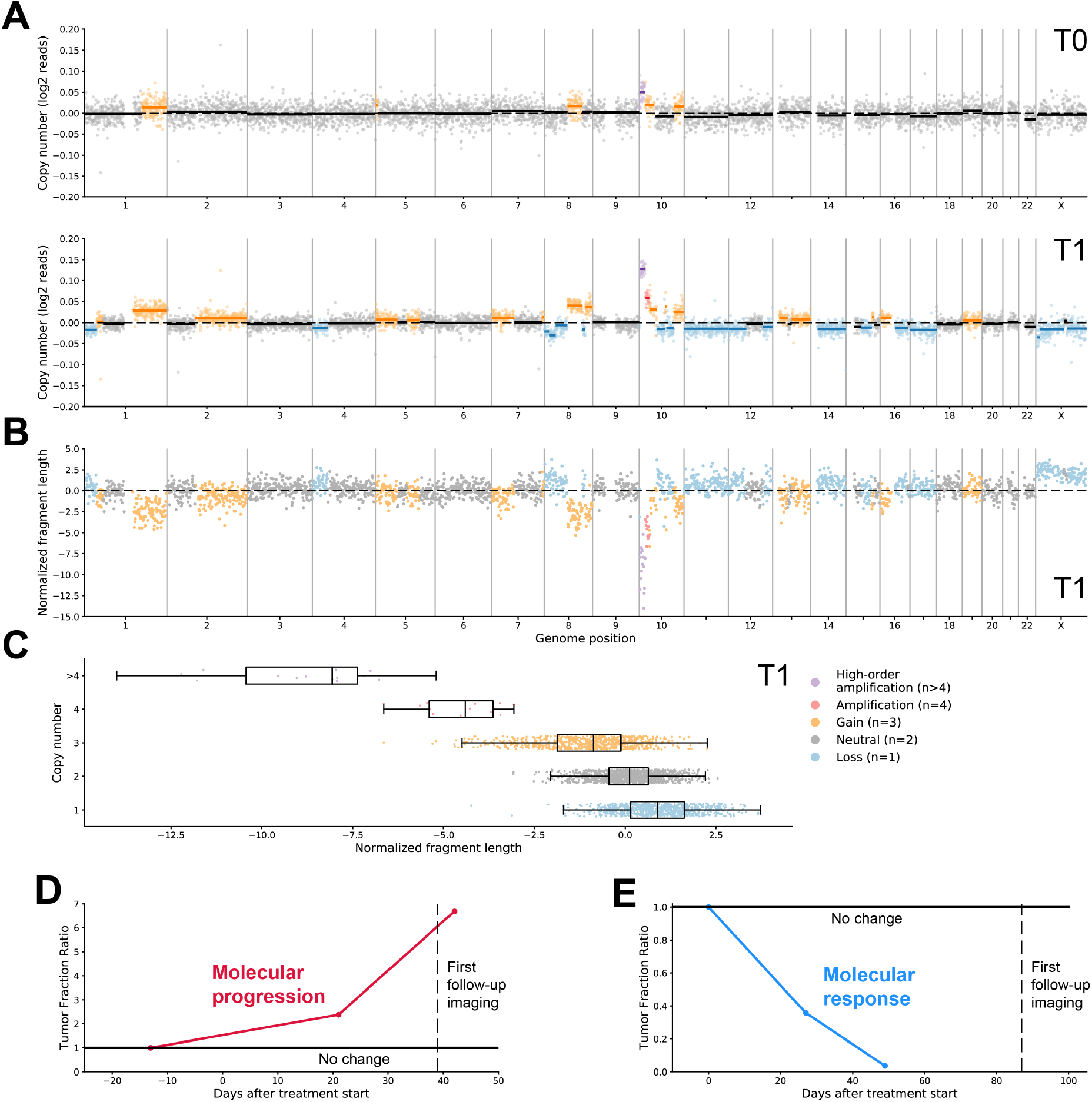
Serial assessment of ctDNA to determine molecular progression. **A**. The genome-wide plots of CNAs detected for patient LS030178. The T0 baseline blood draw was collected 13 days before the start of treatment, and T1 was collected 21 days after the start of treatment. **B**. Normalized fragment length exhibit the reverse pattern compared to CNAs. **C**. Overall, there was a strong negative correlation between the normalized fragment length at each genomic position and the inferred copy number (Spearman’s rho=-0.57, P<10^−10^). **D**. Patient LS030178 had an increase in TFR at follow-up time points T1 and T2, detectable in advance of imaging that indicated progressive disease. **E**. Patient LS030093, who responded to therapy, showed a marked decrease in TFR at T1 and T2, concordant with later imaging that showed a partial response.

A subset of samples had both WGS and WGBS and were used to test whether TFR could be quantified equivalently across sequencing protocols (Supp. Fig. S3). TFR values were highly concordant between WGS and WGBS, enabling analysis of the full cohort including samples analyzed with both protocols.

### Changes in tumor fraction at early time points predict radiographic progression

At FFUI, 24 of 91 patients had PD and 67 had nonPD. To evaluate the predictive value of the ctDNA assay, we compared the classification of MP to FFUI for all patients (Fig. 3A). All 13 patients with an MP at either T1 or T2 had PD, and of the 66 patients who were nonPD, none had MP. The sensitivity of the assay including time points T1 and T2 was 54% and specificity was 100%. The sensitivity was 41% at T1 and was 65% at T2 (Fig. 3B). While the sensitivity at individual time points increased between T1 and T2, there was no statistically significant relationship between sensitivity and timing of the blood draw (Kolmogorov-Smirnov test P=0.077, Supp. Fig. S4). In cases where MP was called, the time point where it was first identified preceded the detection of progression by imaging as scheduled by standard practice by a median of 39 days (Fig. 3C).

**Figure 3.**
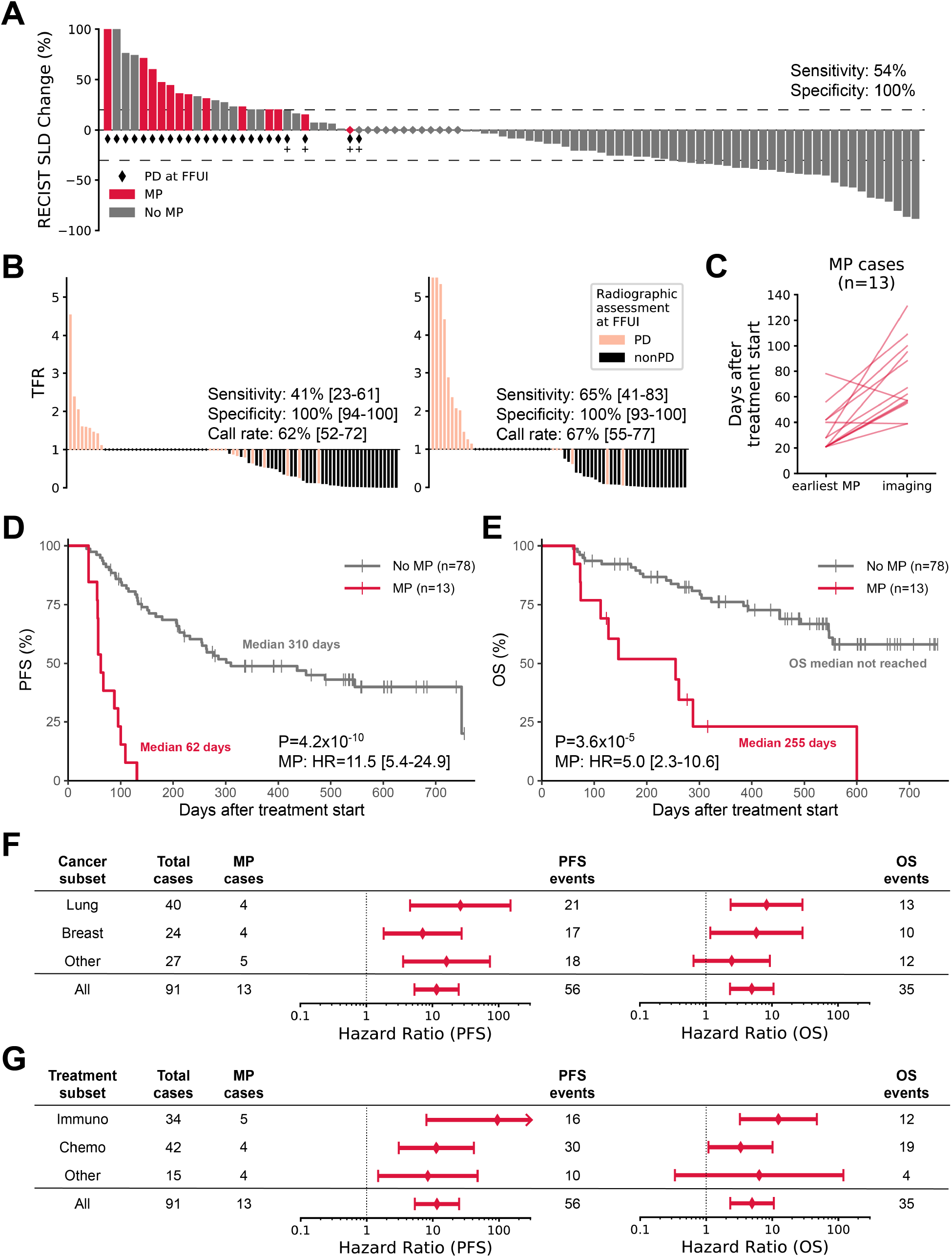
ctDNA assessments following first or second cycle of therapy predicted progression. **A**. Comparison of imaging results at FFUI, sum of longest diameters (SLD) assessed by RECIST 1.1, with ctDNA assessment of molecular progression, indicated by a confident increase in TFR for either post-treatment sample (n=91, sensitivity=54%, specificity=100%). Footnoted cases, designated with a plus sign, showed clear clinical progression. **B**. TFR for progressors and non-progressors at T1 (left, n=85) and T2 (right, n=66), compared to radiographic or clinical assessment of PD or nonPD, showing predictive performance at each time point. Diamonds indicate no change. **C**. For patients with molecular progression (n=13), detection of the molecular progression was observed to precede the date of detection of progression by standard of care imaging by a median of 39 days. Two cases showed molecular progression after FFUI, by 1 day and 21 days. **D-E**. PFS **(D)** and OS **(E)** plotted for all patients grouped by molecular progression. Patients with MP had significantly shorter PFS (P=4.2×10^−10^) and OS (P=3.6×10^−5^). **F-G**. Hazard ratio with 95% confidence intervals for **(F)** cancer type subsets and **(G)** treatment modality subsets. Other cancer types include gastrointestinal, genitourinary, melanoma, and sarcoma. Other treatment modalities include endocrine and/or targeted therapy.

Among the 11 patients with PD at FFUI for whom MP was not called on either time point, three had no confident CNAs detected, two had no significant change in tumor fraction, and six had a decrease in tumor fraction. For the discordant cases with a decrease in tumor fraction, there were a variety of cancer types represented (three breast, one lung, one renal, one sarcoma), treatments (three chemotherapy, one immunotherapy, one endocrine therapy, one targeted therapy alone), and lines of therapy (two first, two second, one third, one fifth). Additionally, there were no major differences in predictive performance between WGS and WGBS (Supp. Table S5).

### Molecular response assessment correlates with PFS and OS

We examined PFS and OS to evaluate the predictive value of early molecular response assessment. The full cohort of patients (n=91) had median PFS of 255 days and median OS of 600 days (Supp. Fig. S5). Patients with MP at either T1 or T2 (n=13) had significantly shorter PFS (HR=11.5 [95% CI 5.4-24.9], P=4.2×10-10), with a median PFS of 62 days vs. 310 days for patients with no MP (n=78; Fig. 3D). These patients also had significantly shorter OS (HR=5.0 [95% CI 2.3-10.6], P=3.6×10-5), with 255 days median OS for patients with MP (n=13) and OS median not reached for those with no MP (n=78; Fig. 3E).

Clinical covariates may also contribute to survival in our heterogeneous cohort. In univariate analysis (Supp. Table S6) prior treatment was the only clinical variable associated with PFS (P=0.0038), and none of the clinical covariates were significantly associated with OS. In multivariable analysis, we found that after controlling for line of therapy (Supp. Table S7), MP remained strongly associated with both PFS and OS.

We explored the predictive performance in subgroups based on tumor origin (Fig. 3F, Supp. Fig. S6) and treatment type (Fig. 3G, Supp. Fig. S7). Cancer type subgroups included lung cancer (n=40), breast cancer (n=24), and the remainder of mixed tumor types (n=27) including GI, GU, melanoma, and sarcoma. We found that MP was significantly associated with both PFS and OS in both lung cancer and breast cancer subsets, and with PFS but not OS in the remaining cases. Subgroups based on the type of systemic therapy received included immunotherapy (with or without chemotherapy or histone deacetylase inhibitor, n=34), chemotherapy (with or without targeted or endocrine therapy, n=42) and other patients (n=15), who were on targeted and/or endocrine therapy alone. While MP was significantly associated with OS for the immunotherapy and chemotherapy subgroups, the association did not reach significance for the smaller subgroup of endocrine and/or targeted therapy. However, MP was significantly associated with PFS for all treatment type subgroups. Additionally, we found that MP was significantly associated with both PFS and OS for subsets of patients who were treatment naive or had prior lines of treatment (Supp. Fig. S8). Predictions of radiographic progression also had comparable performance across these subsets (Supp. Table S8).

### Major molecular response early on treatment predicts long-term outcome

We hypothesized that a large quantitative reduction early in the treatment course would be associated with an improved outcome. Of the 78 patients with no MP, 27 had an MMR at either post-treatment time point, defined as a 10-fold decrease in TFR. Notably, in all cases where an MMR was identified at T1, this finding was also observed at T2, if that time point was available (n=12). In seven cases the TFR from baseline did not reach a 10-fold reduction at T1 but did at T2, for example for patient LS030093 (Fig. 2E).

We found that patients with an MMR (n=27) had longer PFS (Fig. 4A, median PFS of 749 days) compared to patients with no MP and no MMR (n=51; median PFS of 211 days), a difference that was significant (MMR: HR=0.45 [95% CI 0.22-0.92], P=0.028, Supp. Table S9). These patients also had significantly longer OS (Fig. 4B; MMR: OS median not reached, HR=0.116 [95% CI 0.027-0.496], P=0.0036) compared to patients with no MP and no MMR (median OS of 546 days). A multivariable analysis controlling for line of therapy (Supp. Table S10) showed that MMR remained strongly associated with OS. However, the association was marginal for PFS.

**Figure 4.**
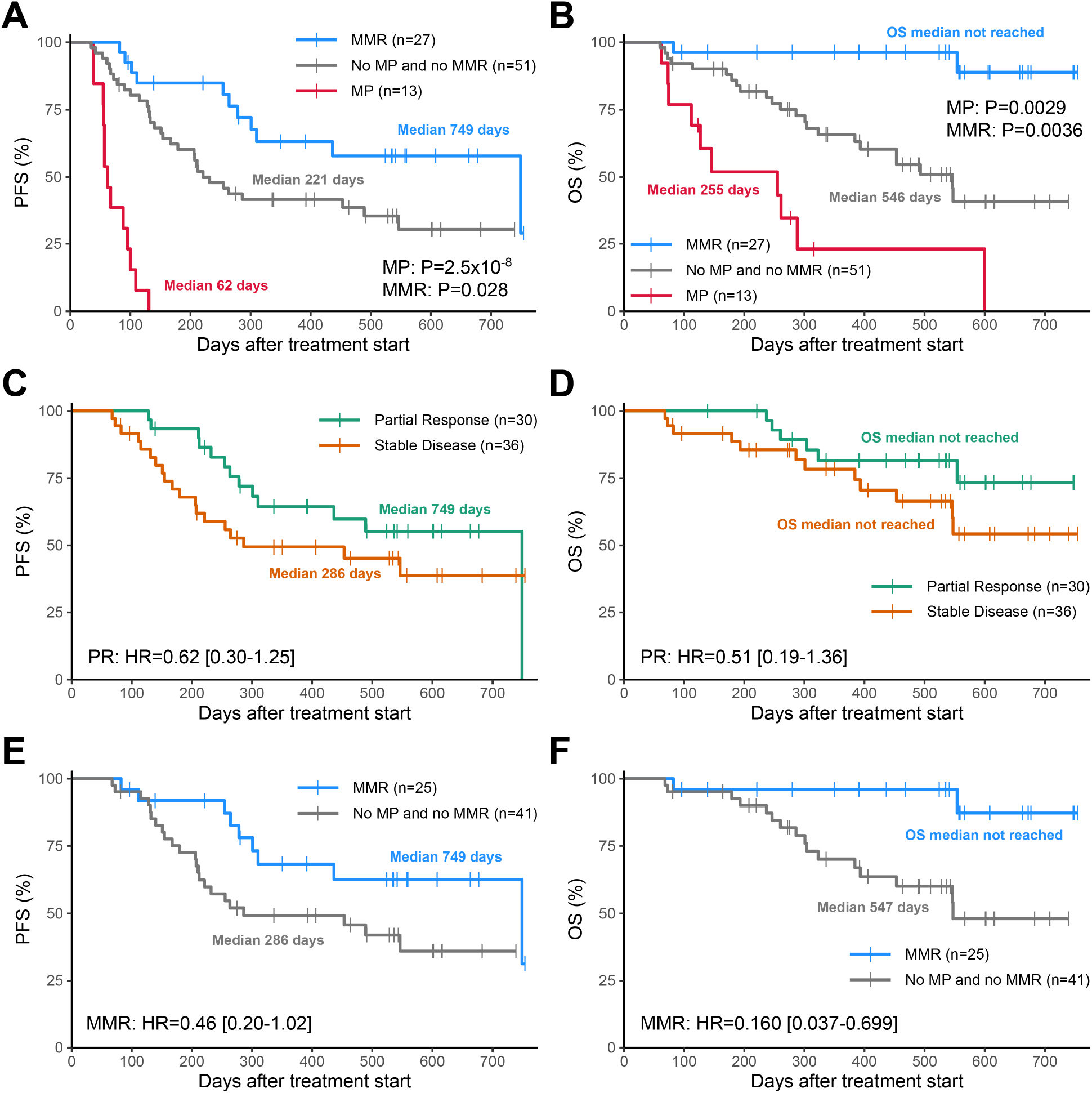
MMR was associated with a favorable outcome. **A-B.** PFS **(A)** and OS **(B)** for patients with MP or MMR. Patients with MMR had significantly longer PFS (P=0.028) and OS (P=0.0036). **C-F**. Survival analysis of the subset of patients with nonPD assessed radiographically at FFUI (N=66), stratified by **(C-D)** response status at FFUI or **(E-F)** MMR.

Next, we sought to assess the predictive value of an MMR in the context of radiographic response monitoring. For patients with radiographic PR or SD at FFUI (n=66), the radiographic assessment of PR or SD had limited prognostic value for PFS (Fig. 4C; HR=0.62 [95% CI 0.30-1.30]) or OS (Fig. 4D; HR 0.50 [95% CI 0.19-1.36]). Among the same subset, patients with an MMR (n=25) had substantially longer PFS (HR=0.46 [95% CI 0.20-1.02]) and OS (HR=0.160 [95% CI 0.037-0.699]) than patients with no MP and no MMR (n=41; Fig. 4E-F, Supp. Fig. S9). These results indicate that early quantitative ctDNA dynamics have value for predicting long term treatment efficacy, beyond what is known from initial radiographic assessment.

### Longitudinal changes in methylation levels may complement tumor fraction changes

To assess the potential of methylation changes for identifying early response to therapy, the change in genome-wide methylation levels from baseline to post-treatment was examined retrospectively for two example patients (Fig. 5)—one with a nonPD call (LS030083) at FFUI and one with a PD call at FFUI (LS030078). Consistent with the clinical assessment at FFUI, a marked increase in methylation levels was observed for LS030083 (Fig. 5A), while a decrease was observed in LS030078 (Fig. 5B). For patient LS030078, a clear molecular progression was observed based on CNAs and local fragmentation changes (TFR=2.01), while no CNAs were detectable in LS030083.

**Figure 5.**
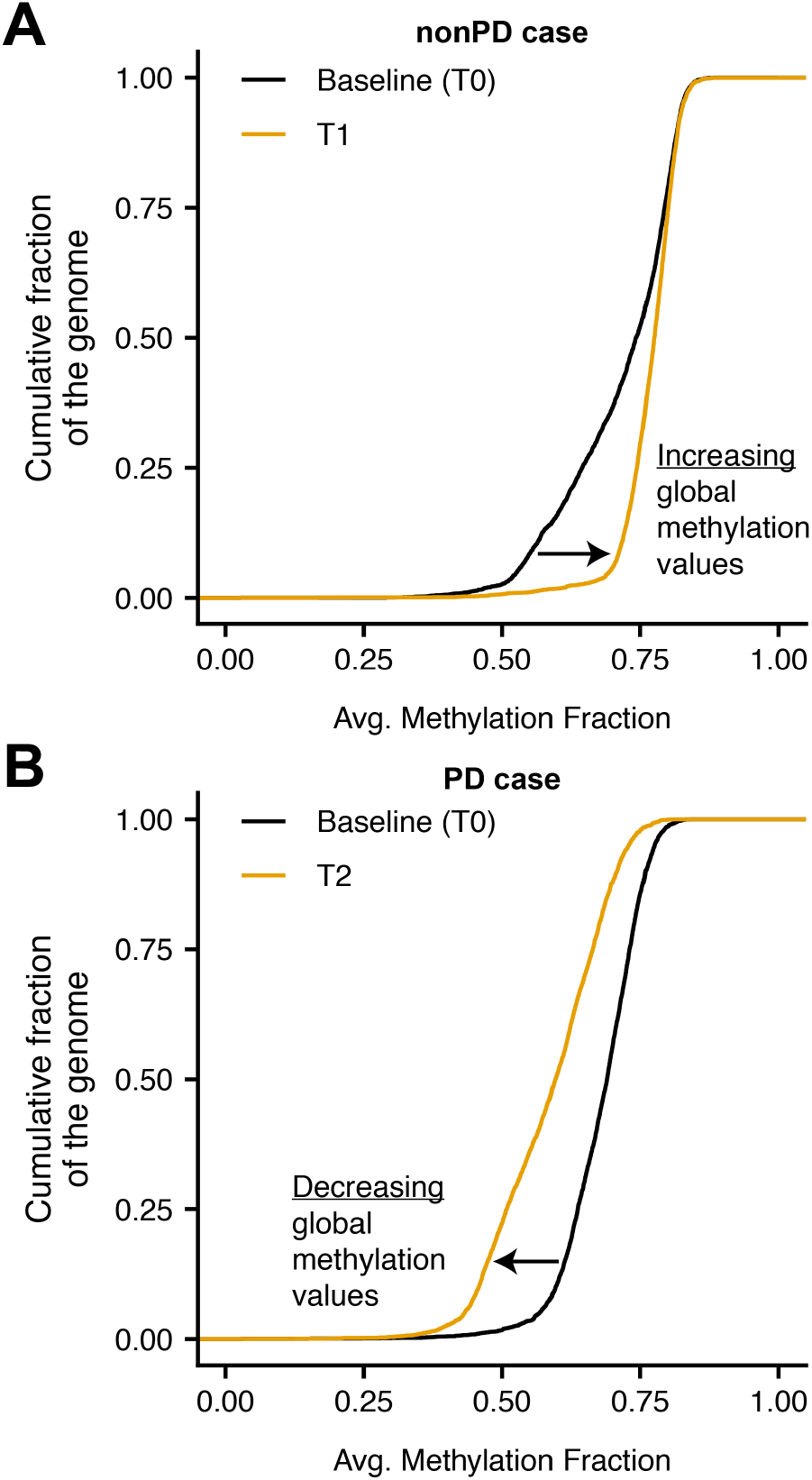
Methylation may provide an orthogonal signal to CNAs for response monitoring. **A-B.** Distribution of average methylation levels in genome-wide 1 megabase bins for **(A)** LS030083, a patient with nonPD, and **(B)** LS030078, a patient with PD. Distributions are plotted at baseline (black line) and during treatment (orange line).

## Discussion

Patients with advanced malignancies require careful treatment monitoring to assess therapeutic efficacy, promote quality of life, and limit drug toxicity. Current methods for disease monitoring using clinical and radiographic assessment often require several months to confidently determine treatment response. Learning this result with high predictive value at an earlier date would be a major improvement in the management of advanced cancers by significantly accelerating the feedback loop regarding therapy effectiveness.

Here, we assessed the utility of a novel, whole-genome cfDNA molecular response assay, which analyzed longitudinal ctDNA measurements at baseline and during the initial cycles of treatment to identify response to therapy. In addition to showing a strong association with survival as observed in previous studies, this technique identified disease progression with a specificity of 100%, so there was high confidence and reliability for these calls. Moreover, MP calls were performed at a median of 6 weeks before clinical or radiographic methods confirmed these assessments. A number of prior studies have evaluated longitudinal ctDNA dynamics to assess tumor response to systemic therapy. Several of these have found strong concordance with survival endpoints (Diehl et al. 2008; Tie et al. 2015; Pécuchet et al. 2016; Murtaza et al. 2013). Additionally, ctDNA biomarkers have been shown to be associated with outcome assessed by standard of care imaging (Tie et al. 2015; Pécuchet et al. 2016; Jensen et al. 2019). Together, these and our findings indicate that ctDNA changes in the blood reflect treatment response and resistance early in the course of treatment, likely before changes in the shape, density, or size of the target lesions on imaging.

Further, we demonstrated that patients with an MMR had a longer PFS and OS compared to those with no change or a smaller decrease in TFR, indicating that there was quantitative value in the degree of initial response to therapy. The additional prognostic value of identifying an MMR supports the potential to integrate imaging and analysis of serial cfDNA samples to provide an early indication of an extended duration of disease control.

We have also found that the blood-based, WG ctDNA molecular assay detected disease progression consistently across a variety of tumors, in the context of any systemic therapy, by looking beyond targeted assessment of tumor-specific point mutations and fusions. Specifically, subset analyses of the two cohorts with the largest number of patients by cancer type (lung and breast cancer) and by treatment modality (chemotherapy and immunotherapy) demonstrated a strong association between MP and PFS and OS. For other subgroups of cancer site and treatment type, MP was only associated with PFS but not OS. These groups were more heterogeneous and had smaller sample sizes compared to breast and lung cancers and chemotherapy and immunotherapy groups. Further studies based on more homogeneous cohorts with appropriate sample sizes are needed to validate the utility of this assay in each of these clinical settings.

Many prior studies have focused on point mutations, tracked by ultra high depth NGS or digital PCR (dPCR). A challenge for these methods is that they must reliably select driver or truncal mutations in each individual case, a process that may need to be calibrated for cancer and treatment type. A common approach is based on tumor tissue sequencing to identify mutations for tracking, but access to adequate tissue varies by tumor type and may be limiting in an advanced disease setting. Biopsies at this stage are often difficult to perform and run the risk of missing relevant clonal mutations, given the genetic heterogeneity of metastatic lesions. In contrast, low-coverage WG approaches can be used to detect (Adalsteinsson et al. 2017; Mouliere et al. 2018) and track changes (Jensen et al. 2019) at low cost based on blood samples alone. A single assay with consistent performance across multiple cancer types and treatment modalities, as we have observed in this study, could facilitate the broad application of a molecular approach to treatment monitoring.

This study has several limitations. First, despite this being a prospectively enrolled study, there were a number of protocol deviations reflecting a wide range of clinical practice patterns. About 30% of participants were not properly enrolled or followed. The study was purposefully designed not to disrupt the flow of clinical care. We made every effort to include participants who met the inclusion criteria and had blood collections from at least two timepoints to avoid introducing bias.

Second, while the specificity of the assay was very high, which is the critical performance metric for clinical utility in the advanced setting, sensitivity to identify clinical progression early in the treatment course was relatively low. Sensitivity, particularly at the earliest time point, may be improved by including other features such as cancer-associated epigenetic signals. For example, in patient LS030083, there was a marked increase in genome-wide methylation levels from baseline to post-treatment, consistent with a nonPD call at FFUI, but no CNAs were detected (Figure 5A). In future work, we plan to incorporate these methylation-based signals into the assay, along with fragment length and copy number information with the goal of increasing assay sensitivity. However, even with additional orthogonal signals, there will still be a residual false negative rate from a) tumors that do not generate sufficient ctDNA (i.e. non-shedding tumors), b) tumors that have not yet progressed during the earliest cycles of treatment and, c) tumors in which molecular progression by ctDNA analysis and imaging are not in agreement.

Third, the study was not powered to conduct subgroup analysis. However, since there was a significant main effect, we explored the assay’s predictive ability across different cancer and treatment types. We hypothesized that the dynamics of early changes in cancer-associated signals in the blood identified by WGS are agnostic to cancer type as the results of this study suggest. Prospective validation studies are planned to confirm the observed findings to give clinicians the confidence to use this assay to manage treatment.

Molecular response monitoring with serial measurements of cfDNA has potential clinical benefits for both assessing disease progression and disease control, if further validated in an adequately powered prospective clinical validation study. An early call of MP would lead oncologists to consider discontinuation of ineffective treatments, thereby reducing avoidable side effects and financial toxicity. By accelerating the clinical feedback loop, patients would be afforded the opportunity to change to an alternative, potentially effective therapy. In contrast, MMR would provide confidence in the current treatment plan, and encourage clinicians and patients to maintain the course. Clinicians could also potentially reduce the frequency of imaging in these cases. In addition, new adaptive approaches should be considered with MMR as the interim endpoint in clinical trials. Moreover, a blood-based assay provides convenience for patients as blood samples are collected routinely during the course of therapy. Patients may have improved outcomes by limiting side effects and costs associated with ineffective treatments, trying alternate potentially effective treatments earlier, and feeling encouraged with early exceptional response to maintain the course.

In conclusion, we demonstrated that identifying disease progression through earlier low-pass WG ctDNA-based monitoring was concordant with standard of care clinical assessment and the degree of ctDNA reduction was strongly correlated with long-term outcome. These findings were consistent across multiple tumor and treatment types. The assay captured changes in tumor biology that occurred early in the course of systemic therapy, which may be observed before confident radiographic detection of these changes is possible. If validated prospectively, this non-invasive tool would allow oncologists and patients to understand response and resistance to treatment in a more precise and timely way.

## Data Availability

Data in this manuscript is represented in the methods, results, and supplementary information.

## Acknowledgements

We thank all participants and their families for participating in this study. We also thank Dr. Brian McNamee for review of all clinical images, as well as all clinicians and their research staff.

